# Changes in Pediatric RSV Hospitalizations after the COVID-19 Pandemic, 2022-2023, Canada: An Active Surveillance Study

**DOI:** 10.1101/2025.08.13.25333142

**Authors:** Aariana Lopes, Joanne Embree, Taj Jadavji, Kescha Kazmi, Joanne M. Langley, Marc H. Lebel, Nicole Le Saux, Dorothy Moore, Shaun K. Morris, Jeffrey M. Pernica, Joan Robinson, Manish Sadarangani, Hennady Shulha, Julie A. Bettinger, Jesse Papenburg, the Canadian Immunization Monitoring Program Active (IMPACT) Investigators

## Abstract

**Background:** The COVID-19 pandemic impacted RSV epidemiology. We describe Canadian pediatric tertiary care RSV hospitalizations in 2022-2023 and assess pandemic-related changes.

**Methods:** Active surveillance of hospitalized children aged 0-16 years was conducted at 13 Immunization Monitoring Program, Active (IMPACT) centres. RSV hospitalizations in 2022-2023 were compared to those in the pre-pandemic period (2017-2018 through 2019-2020). Province-specific and age-stratified proportions of all-cause hospitalizations with RSV detection and age-stratified proportions of RSV-associated intensive care unit (ICU) admission were calculated. Changes in seasonality were assessed using Seasonal Autoregressive Integrated Moving Average (SARIMA) modelling.

**Results:** In 2022-2023, there were 5362 RSV-associated hospitalizations including 1260 (23.5%) ICU admissions, both more than double pre-pandemic yearly averages. Overall, median (IQR) age increased from 6 months (1-20) to 9 months (2-27; P<0.001). The proportion of RSV hospitalizations among all-cause hospitalizations increased by 3.5 percentage points (95% CI 3.3-3.7 percentage points), to 6.8% (95% CI 6.6%-7.0%). While 41.5% of RSV hospitalizations were among children <6 months old, they accounted for 62% of ICU admissions. Overall, ICU proportion remained constant; however, odds of ICU admission among infants <6 months old increased (adjusted OR 1.35, 95% CI 1.2-1.52) compared to the pre-pandemic period. National weekly incidence in 2022-2023 peaked earlier, higher and persisted longer than expected by SARIMA.

**Interpretations:** In 2022-2023, the number of RSV hospitalizations and ICU admissions increased dramatically in Canadian pediatric hospitals. Despite an older age distribution, the greatest burden remained in children <6 months old. RSV immunization strategies for young infants will likely have substantial potential public health impact.

## Introduction

Respiratory syncytial virus (RSV) is the leading cause of pediatric lower respiratory tract infections, with the highest hospitalization risk in the first months of life.^(1–4)^ Although underlying health conditions predispose to severe RSV illness, healthy children born at term account for most pediatric RSV-associated hospitalizations.^(4–7)^

Prior to the onset of the COVID-19 pandemic, RSV incidence followed consistent seasonal trends in Canada, with activity peaking yearly during the winter.^(8–12)^ The public health interventions implemented in 2020 to counter the COVID-19 pandemic contributed to a marked reduction in respiratory virus transmission.^(13, 14)^ Lack of exposure to RSV at the population level for this prolonged period likely led to an accumulation of susceptible individuals, which may have been the primary cause of the surge in RSV activity in 2021- 2022 when the stringency of public health interventions was reduced. ^(5, 8–10, 15–19)^

Using national active surveillance from pediatric tertiary care centers in the Canadian Immunization Monitoring Program Active (IMPACT) program, we previously demonstrated that, after a near absence in 2020-2021, RSV admissions were higher in 2021-2022 compared to pre-pandemic averages. While severity and age distribution remained similar, there was greater interregional variability in timing of RSV activity.^(15)^ These changes in the timing of RSV circulation, with earlier peaks starting in summer months, have also been observed in the first epidemics since the COVID-19 pandemic in other countries with a temperate climate.^(5, 15, 20–22)^ It is unknown if these trends will persist into future seasons.

There have also been conflicting reports regarding changes in the age distribution and severity of RSV hospitalizations in children since the onset of the pandemic.^(8, 21, 23)^In the late summer and fall of 2022, Canadian healthcare practitioners, public health units and lay media reported a surge in RSV cases, overwhelming pediatric hospital capacity.^(24, 25)^However, national data on pediatric RSV hospitalizations in 2022-2023, including possible interregional differences, are lacking. Understanding these potential changes in epidemiologic characteristics is important to inform pediatric healthcare capacity planning and evidence-based strategies for preventing severe RSV disease such as long-acting monoclonal antibody immunoprophylaxis and RSV vaccination during pregnancy. These immunization products were authorized for use in Canada in 2023 and 2024, respectively.^(26, 27)^ We aimed to describe the overall, age-specific, and region-specific characteristics and in-hospital severity and outcomes of Canadian pediatric tertiary-care RSV hospitalizations in 2022-2023 compared to 3 pre-pandemic seasons (2017-2018 to 2019-2020), using national active surveillance data.

## Methods

Institutional ethics or hospital approval was obtained at each IMPACT centre for this active surveillance study, with waiver of informed consent due to the minimal risk of data collection associated with this health record review. This study follows the Strengthening the Reporting of Observational Studies in Epidemiology (STROBE) reporting guideline for observational studies.^(28)^

### Study Population, Design and Setting

We performed active surveillance for all laboratory-confirmed hospitalized cases of RSV infection in IMPACT hospitals among children 0-16 years of age. Surveillance was year-round during the 3 most recent seasons (July 2020 to June 2023) and seasonal during the pre-pandemic seasons (October 2017 to June 2018, November 2018 to June 2019, and October 2019 to June 2020). Twelve hospitals participated during the pre-pandemic period. An additional center was added in July 2020. Data from this site were excluded from all statistical comparisons between time periods. Participating IMPACT hospitals account for approximately 90% of the pediatric tertiary care beds in Canada and admit patients from all provinces and territories.^(29, 30)^

### Data Collection

Trained nurse monitors identified RSV cases by screening laboratory results and admission lists. Data collection included hospital admission and discharge dates, age, sex, RSV specimen collection date, ICU admission, and outcome at discharge or transfer (died or survived). We also captured the total number of all-cause hospitalizations for each year from each IMPACT hospital (excluding psychiatry wards and newborn nurseries). Data on underlying medical conditions were not collected. We could not distinguish between hospital-acquired and community-acquired infections. Data were recorded after patient discharge from medical records using an electronic case report form with data validation checks to improve completeness and accuracy.

### Statistical Analysis

We considered 3 RSV seasons (2017-2018, 2018-2019, and 2019-2020) as the pre-pandemic period. Data from 2020-2021 and 2021-2022 were excluded from statistical analyses to focus comparisons between 2022-2023 and the pre-pandemic seasons. The Wilcoxon test was used for comparison of median age (in months), while categorical variables were compared using the χ^2^ test. P values ≤0.05 were considered statistically significant, and all comparisons were 2-sided.

The overall and province-specific proportions and their 95% confidence intervals (CI) of all-cause hospitalizations with RSV detection in 2022-2023 were compared with the pre-pandemic seasons using a two independent groups proportion test to calculate percentage point difference. Severity markers for RSV hospitalizations, including ICU admission (the primary severity outcome), length of stay ≥ 7 days, and death, also using a two independent groups proportion test. Subgroup analyses were conducted to study disease severity differences by age group (i.e. 0-5 months, 6-11 months, 12-23 months, 2-4 years, 5-9 years, and 10-16 years). A mixed effects logistic regression model, adjusting for sex and using IMPACT site as a random effect, assessed the association between ICU admission in 2022- 2023 versus the pre-pandemic period, stratified by age group.

A weekly epidemic curve for 2022-2023 was plotted with the pre-pandemic average and 2021-2022 overlaid. Monthly epidemic curves were plotted for each province and the pre-pandemic average. Seasonal autoregressive integrated moving average (SARIMA) modelling was used to compare the expected weekly case counts, using pre-pandemic trends, to the observed 2022-2023 weekly case counts.

Statistical analyses were performed using R statistical software version 4.3.3. Bonferroni corrections were applied to P-values for multiple statistical comparisons.

## Results

### Age and Sex Distributions

In 2022-2023, 5872 RSV hospitalizations occurred in the 13 IMPACT hospitals. They included 2625 (44.7%) females and 2425 (41.3%) children aged 0-5 months. Sex distribution was consistent with the pre-pandemic period. All age groups experienced approximately two to three times the number of cases compared to pre-pandemic yearly totals; however, there were changes in the age distribution **(Table 1)**. Median age increased from 6 months (interquartile range [IQR] 1-20) to 9 months (IQR 2-27; P<0.001). Children aged 0-5 months accounted for a smaller proportion of total cases (41.5% in 2022-2023 vs. 48.9% in the pre-pandemic seasons [corrected P<0.001]). In contrast, there was an increase in the proportion of cases among children aged 2-4 years (21.3% in 2022-2023 vs. 15.5% in the pre-pandemic seasons [corrected P<0.001] and 5-9 years (5.7% in 2022-2023 vs. 3.9 % in the pre-pandemic seasons [corrected P<0.001]).

**Table 1.** Age and Sex Distribution of Laboratory Confirmed Respiratory Syncytial Virus (RSV) Hospitalizations, Canadian Immunization Monitoring Program, ACTive.

| **RSV hospitalizations, No. (%)<sup>a</sup>** |  |  |  |
| --- | --- | --- | --- |
| **Characteristic** | **2017-2018 to 2019-2020 (N=7549)** | **2022-2023<sup>b</sup> (N=5362)** | **P-value<sup>c</sup>** |
| **Age (months)** |  |  |  |
| Mean (SD) | 16.4 (27.6) | 19.8 (28.1) |  |
| Median (IQR) | 6 (1-20) | 9 (2-27) | <0.001 |
| **Sex<sup>d</sup>** |  |  |  |
| Female | 3424 (45.4) | 2398 (44.7) | 0.477 |
| Male | 4124 (54.6) | 2963 (55.3) |  |
| **Age group** |  |  |  |
| 0-5 months | 3693 (48.9) | 2225 (41.5) | <0.001 |
| 6-11 months | 928 (12.3) | 677 (12.6) | 1 |
| 12-23 months | 1330 (17.6) | 921 (17.2) | 1 |
| 2-4 years | 1170 (15.5) | 1140 (21.3) | <0.001 |
| 5-9 years | 292 (3.9) | 303 (5.7) | <0.001 |
| 10-16 years | 136 (1.8) | 96 (1.8) | 1 |

<sup>a</sup> 2020-2021 and 2021-2022 seasons are excluded from all analyses in this table.
<sup>b</sup> An additional Ontario site participated as of July 1, 2020. Data from this site were excluded from this analysis and are not included in table totals.
<sup>c</sup> Bonferroni corrected p-values are presented for age groups.
<sup>d</sup> There was 1 case with value for sex missing.

### RSV Hospitalizations Among All-Cause Hospitalizations

Compared to the pre-pandemic annual average of 2517, RSV hospitalization counts more than doubled in 2022-2023 to 5362, which represented an increase of 3.5 percentage points (95% CI 3.3-3.7 percentage points, corrected P<0.001) in the percentage of all-cause hospitalizations in which RSV was detected (6.8% in 2022-2023 vs 3.3% in 2017- 2018 to 2019-2020). All provinces except Nova Scotia experienced a significant increase in the proportion of RSV hospitalizations among all-cause hospitalizations **(Table 2 and Supplemental Table 1)**.

**Table 2.** Proportions of All-Cause Admissions with Respiratory Syncytial Virus (RSV) Detected, Canadian Immunization Monitoring Program, ACTive^a^.

| Province | Proportion of RSV hospitalizations, % [95%CI] |  | Difference [95% CI], percentage points | P-value<sup>b</sup> |
| --- | --- | --- | --- | --- |
|  | 2017-2018 to 2019-2020 | 2022-2023 |  |  |
| Alberta | 3.4 [3.2-3.5] | 9.7 [9.2-10.1] | 6.3 [5.8-6.8] | <0.001 |
| British Columbia | 2.8 [2.5-3] | 6.1 [5.5-6.7] | 3.3 [2.7-4] | <0.001 |
| Manitoba | 4.9 [4.6-5.3] | 9.1 [8.3-9.9] | 4.1 [3.3-5] | <0.001 |
| Newfoundland and Labrador | 3.7 [3.2-4.3] | 5.6 [4.5-6.8] | 1.8 [0.6-3.1] | 0.014 |
| Nova Scotia | 4.1 [3.8-4.5] | 4.2 [3.6-4.9] | 0.1 [-0.7-0.8] | 1 |
| Ontario<sup>c</sup> | 2.2 [2.1-2.3] | 4 [3.7-4.3] | 1.8 [1.5-2.1] | <0.001 |
| Quebec | 3.9 [3.7-4] | 7.6 [7.2-8] | 3.7 [3.3-4.1] | <0.001 |
| Saskatchewan | 3.8 [3.4-4.2] | 7.4 [6.5-8.3] | 3.6 [2.6-4.5] | <0.001 |
| **Total** | 3.3 [3.2-3.4] | 6.8 [6.6-7] | 3.5 [3.3-3.7] | <0.001 |

<sup>a</sup> 2020-2021 and 2021-2022 seasons are excluded from all analyses in this table.
<sup>b</sup> Bonferroni corrected p-value
<sup>c</sup> An additional Ontario site participated as of July 1, 2020. Data from this site were excluded from this analysis.

### Disease Severity

In 2022-2023, 1260 (23.5%) RSV hospitalizations required admission to the ICU **(Table 3)** of which 782 (62%) were in children aged 0-5 months **(Supplemental Table 2)**. While the overall proportion of ICU admissions remained constant in 2022-2023 compared to the pre-pandemic seasons, the absolute number was more than double the yearly pre-pandemic average **(Table 3)**. Infants <6 months of age had increased odds of ICU admission in 2022-2023 compared to the pre-pandemic seasons (OR, 1.35, 95% CI 1.2-1.52), while children aged 12-23 months (OR 0.75, 95% CI 0.59-0.95) and 2-4 years (OR, 0.76, 95% CI 0.60-0.96) had decreased odds of ICU admission **(Figure 1)**. The proportion of RSV hospitalizations with a hospital length of stay > 7 days was significantly lower in 2022- 2023 (corrected P=0.008) while mortality was similar to the pre-pandemic seasons **(Table 3)**.

**Figure 1.**
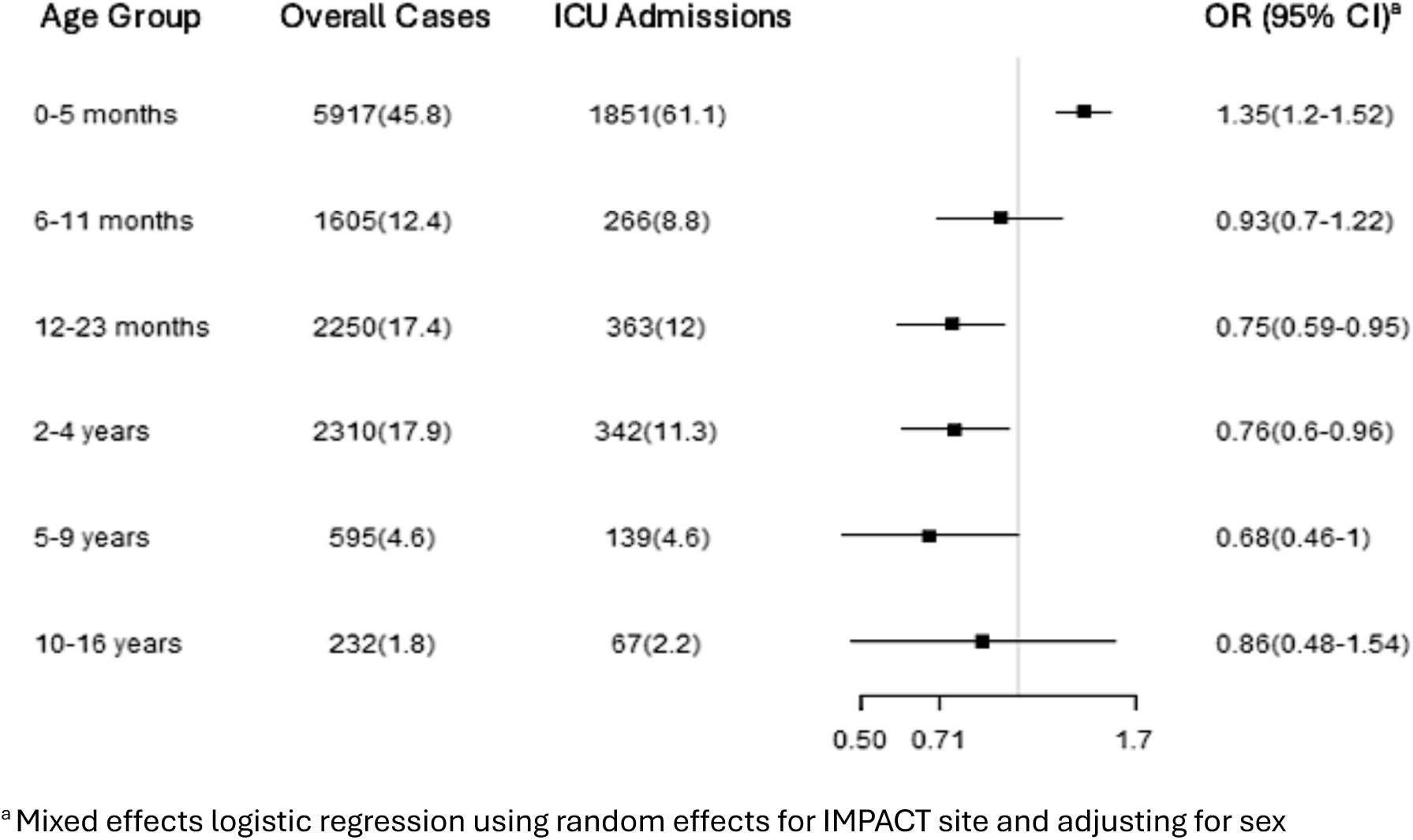
Adjusted odds ratios of ICU admission, stratified by age group, among RSV hospitalizations in 2022-2023 compared to pre-pandemic period (2017-2018 to 2019-2020), Canadian Immunization Monitoring Program, ACTive

**Table 3.**
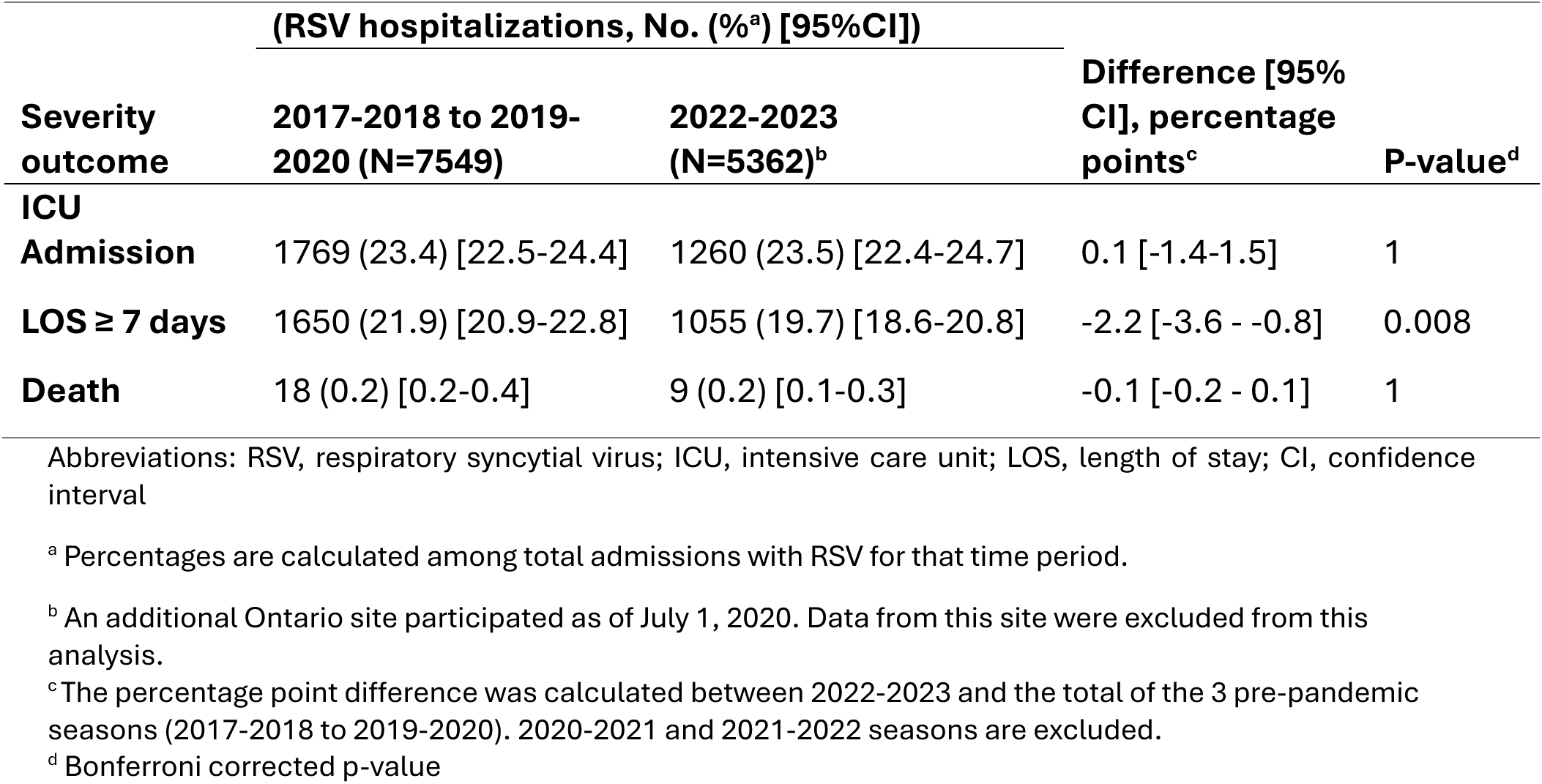
Severity of Respiratory Syncytial Virus (RSV) Hospitalizations, Canadian Immunization Monitoring Program, ACTive.

### Seasonality and Regional Variation

In 2022-2023, nationally, peak weekly RSV hospitalization incidence occurred at epidemiologic week 24 (early December 2022) (**Figure 2).** There was variation between provinces in monthly peaks. Ontario and Quebec peaked earliest, in November, and Saskatchewan had the latest peak, in January **(Figure 3).** The observed national peaks in weekly incidence in 2021-2022 and 2022-2023 differed from the predicted weeks **(Figure 4)**. The peak in 2021-2022 was earlier and lower than predicted, while the peak in 2022- 2023 was earlier, much higher and declined earlier than predicted with a very small number of cases persisting longer than predicted.

**Figure 2.**
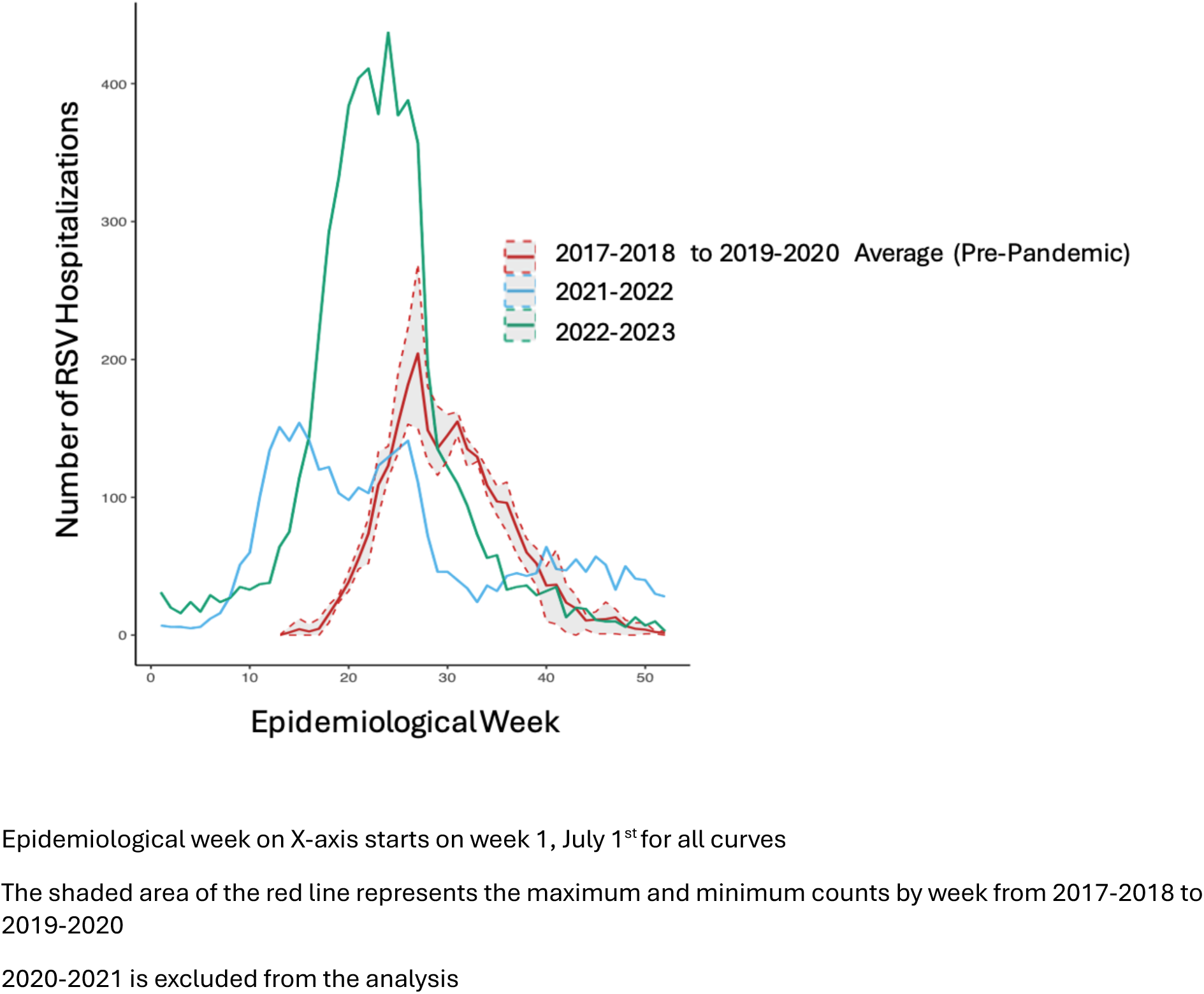
Weekly Respiratory Syncytial Virus (RSV) Hospitalizations, 2017-2022, Canadian Immunization Monitoring Program, ACTive

**Figure 3.**
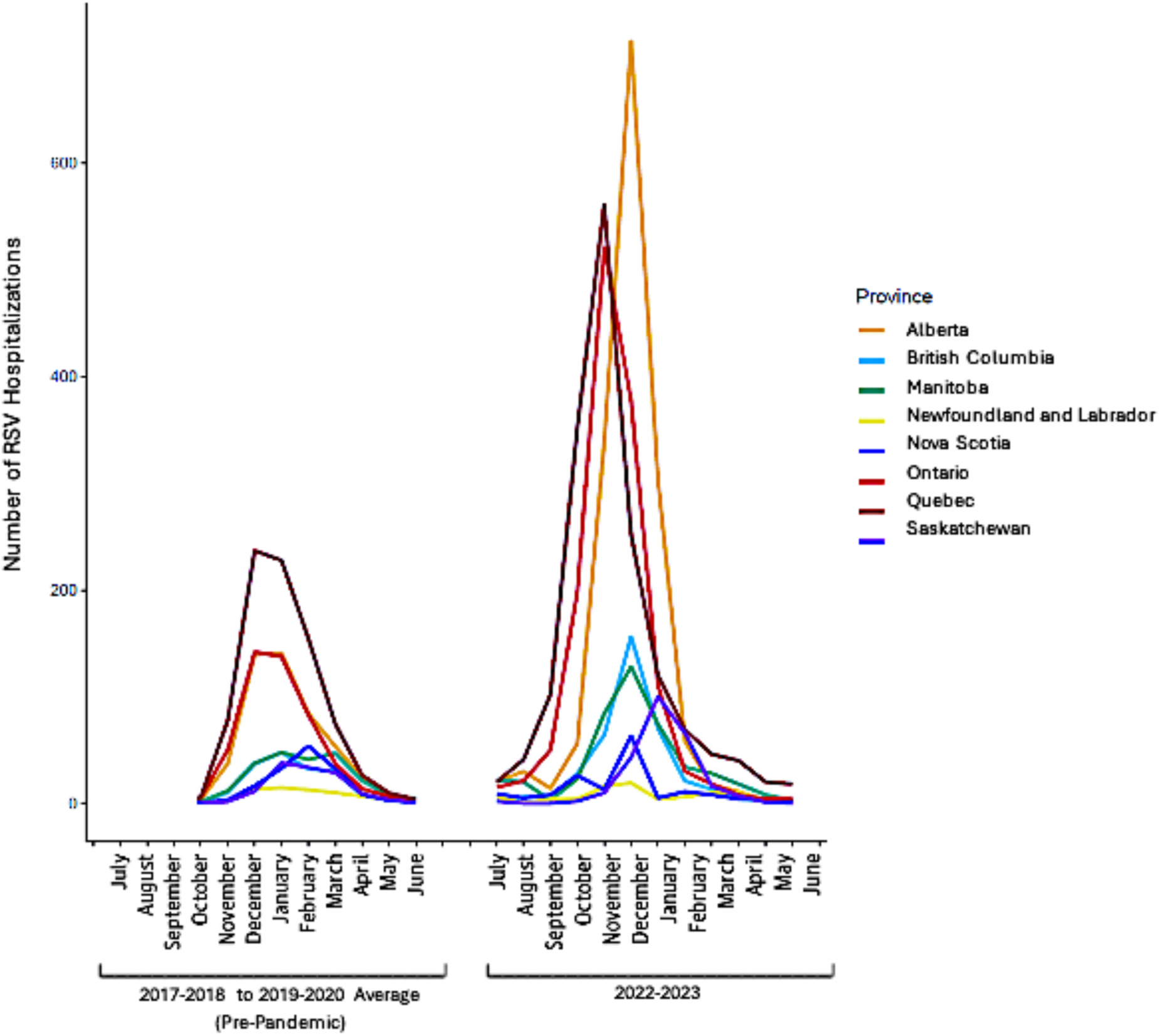
Monthly Respiratory Syncytial Virus (RSV) Hospitalizations, by Province, 2017-2018 to 2022-2023, Canadian Immunization Monitoring Program, ACTive

**Figure 4.**
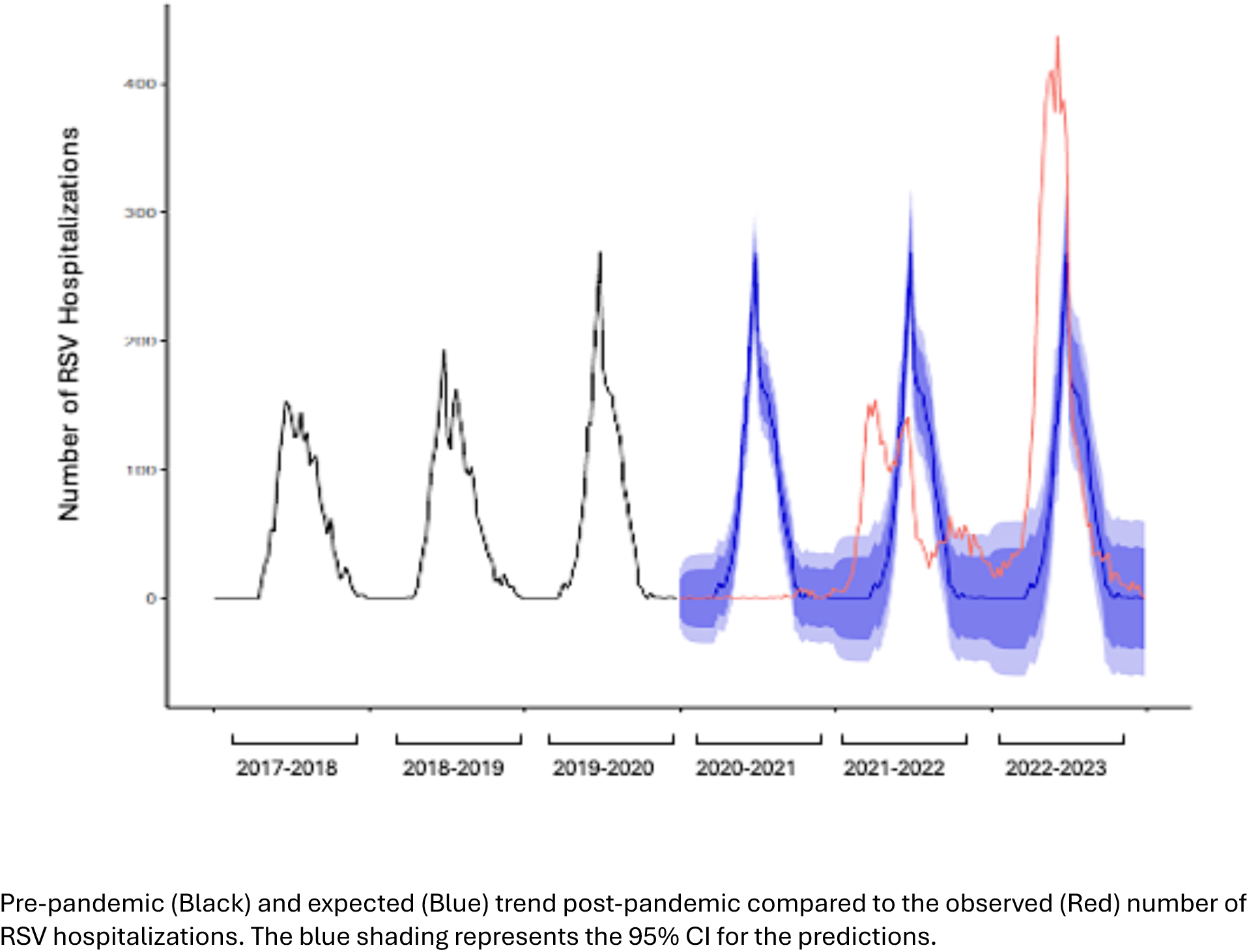
Seasonal Autoregressive Integrated Moving Average (SARIMA) analysis for weekly RSV hospitalizations, 2017-2018 to 2022-2023, Canadian Immunization Monitoring Program, ACTive

### Interpretations

Similar to reports from the United States and Europe, this Canadian national active surveillance study found that the number of pediatric tertiary care RSV hospitalizations and ICU admissions more than doubled in 2022-2023 compared to pre-pandemic yearly averages, exerting a remarkable strain on pediatric hospital capacity during an early and intense RSV season^(21, 22, 31, 32)^. Despite an older age distribution in 2022-2023, the greatest RSV burden in Canadian children unquestionably remained in infants less than 6 months old, with this age group accounting for over 40% and 60% of RSV hospitalizations and ICU admissions, respectively.

Studies of the 2022-2023 RSV season in Ontario, British Columbia, the United States and Europe also saw increases in age distribution.^(5, 8, 23, 31, 32)^ Our study found that a greater proportion of cases were seen in children 2-4 and 5-9 years of age, with both groups experiencing triple their pre-pandemic average number of cases. Similarly, a Danish study found that in 2022-2023, children 3-4 years of age had almost 4 times the risk of hospitalization with RSV compared to the 4 seasons prior to the COVID-19 pandemic.^(21)^ It is notable that in this context we found that children under 6 months of age had 35% increased odds of ICU admission in 2022-2023 compared to the pre-pandemic period, whereas the odds of ICU admission decreased in older age groups. It is likely that young infants, who are most vulnerable to RSV-associated mortality, were given priority to the limited number of pediatric ICU beds. ^(33–35)^

The altered RSV seasonality after the onset of the COVID-19 pandemic and the observed resurgence and older age of affected children in 2022-2023 has been principally attributed to disruptions in transmission patterns caused by strict non-pharmaceutical interventions (i.e. social distancing, mask wearing, school and daycare closures, etc.) at the beginning of the COVID-19 pandemic.^8,9^ This delayed exposure to respiratory viruses, including RSV, resulted in the accumulation of susceptible individuals in the general population, with many children only acquiring their first RSV infection after the first or second year of life once RSV transmission was re-established after the most stringent non-pharmaceutical interventions were lifted.^(7, 9, 19, 20)^ Recent seroepidemiologic studies in Denmark and British Columbia both concluded that there was a population level decrease in RSV antibody-mediated immunity temporally associated with the implementation of COVID-19 pandemic mitigation measures. This included decreased transplacental antibody transfer that passively protects infants in the first months of life.^(18, 19)^ The historically large incidence of RSV infections in the general population in 2022-2023 is likely the cause of infants and older children experiencing higher case numbers of hospitalizations due to the intense community transmission.^(10, 25, 33)^

In 2022-2023, the peak in RSV hospitalizations was earlier and higher than expected. However, our also data show that the national epidemic curve may obscure regional differences. Thus, monitoring the timing of RSV activity at both the national and provincial levels remains essential to optimize the timing of delivery of RSV prevention programs such as long-acting monoclonal antibodies and vaccination during pregnancy.^(36, 37)^

### Limitations

Data collection occurred solely in tertiary pediatric care centers so the results may not be generalizable to the entire Canadian pediatric population. Due to the limited number of variables collected, we could not identify which children had comorbidities, distinguish between community-acquired and hospital-acquired infections, or identify incidental RSV infections unrelated to the indication for hospitalization. Changes in testing practices in 2022-2023 compared to the pre-pandemic period potentially led to an overestimation of the increase in RSV hospitalizations. However, RSV testing among the most severe cases, those admitted to ICU, is unlikely to have changed over the period of surveillance and the observed increase in RSV ICU admissions paralleled the increase in overall RSV hospitalizations in our study and the proportion admitted to ICU remained unchanged. There were gaps in surveillance in the summer months during the pre-pandemic period. However, national laboratory surveillance showed that RSV detections during weeks not covered by IMPACT surveillance accounted for only 0.8% to 1.4% of all RSV detections, yearly.^(38)^ Despite these limitations, this active surveillance study represents approximately 90% of pediatric tertiary care beds in Canada, providing a comprehensive view of RSV hospitalizations during the unique 2022-2023 season and allowing for comparisons over time and across geographic areas.^(29, 30)^

## Conclusion

This national active surveillance study found that in 2022-2023, the number of RSV hospitalizations and ICU admissions increased dramatically in Canadian pediatric hospitals. Despite a shift towards an older age distribution, the greatest burden by far remained in children less than 6 months old. Since their approval in Canada in 2023 and 2024, implementation of highly effective RSV immunization immunoproxylaxis with long- acting monoclonal antibodies and vaccination during pregnancy has varied across provinces.^(27, 39–42)^ Increased and equitable RSV immunization strategies to protect young infants has substantial potential public health impact.

## Data Availability

Data are not available as they belong to the Canadian Paediatric Society

## Author Contributions

Dr Papenburg and Dr Bettinger had full access to all of the data in the study and take responsibility for the integrity of the data and the accuracy of the data analysis.

*Concept and design*: Papenburg, Bettinger

*Acquisition, analysis, or interpretation of data*: Lopes, Shulha, Embree, Jadavji, Kazmi, Langley, Lebel, Le Saux, Moore, Morris, Pernica, Robinson, Sadarangani, Bettinger, Papenburg

*Drafting of the manuscript*: Lopes, Papenburg

*Critical revision of the manuscript for important intellectual content*: Lopes, Embree, Jadavji, Kazmi, Langley, Lebel, Le Saux, Moore, Morris, Pernica, Robinson, Sadarangani, Bettinger, Papenburg

*Statistical analysis*: Shulha

*Obtained funding*: Bettinger, Sadarangani

*Supervision*: Papenburg, Bettinger

## Conflict of Interest Disclosures

Dr. Papenburg reports grants from MedImmune and Merck to his institution, and personal fees from Enanta, all outside the submitted work.

Dr. Sadarangani is supported via a salary award from the BC Children’s Hospital Foundation; in the last 3 years, he has been an investigator on projects funded by GlaxoSmithKline, Merck, Moderna, Pfizer, and Sanofi-Pasteur with all funds paid to his institute, and he has not received any personal payments.

Dr. Halperin has served on ad hoc advisory boards and has been an investigator on projects funded by Sanofi-Pasteur, GlaxoSmithKline, Merck, Pfizer, VBI, CanSino, Moderna, and Seqirus.

Dr. Morris has served on ad-hoc advisory boards for Pfizer, Sanofi Pasteur, GlaxoSmithKline, and Merck and has received speaker fees from GlaxoSmithKline, Sanofi Pasteur, and Pfizer, all unrelated to this study.

Dr. Pernica reports receiving funding (all to McMaster) in grants from AstraZeneca and Merck and in-kind (diagnostics and consumables) in grants from bioMerieux and Abbott.

Dr. Langley has been an investigator on projects funded by GlaxoSmithKline, Merck, Moderna, Pfizer, Sanofi-Pasteur, Seqirus, Symvivo, Entos, VBI Vaccines and CanSino with all funds paid to her employer, Dalhousie University and he has not received any personal payments. Dr. Langley holds the CIHR-GSK Chair in Pediatric Vaccinology at Dalhousie University.

Dr Robinson has received honoraria for presentations from the Alberta Pharmacists’ Association and the Canadian Paediatric Society and received payment for peer review from Elsevier.

Dr. Kazmi has received honoraria for ad-hoc advisory boards for Takeda Canada and Bavarian Nordic, all unrelated to this study.

The other authors have no conflicts of interest to disclose.

## Funding/Support

This surveillance activity is conducted as part of the Canadian Immunization Monitoring Program Active (IMPACT), a national surveillance initiative managed by the Canadian Pediatric Society (CPS) and conducted by the IMPACT network of pediatric investigators on behalf of the Public Health Agency of Canada’s (PHAC’s) Centre for Immunization and Respiratory Infectious Diseases. Funding for RSV surveillance was provided by PHAC. Participation by Aariana Lopes was supported by Mitacs through the Mitacs Accelerate program

## Role of Funder/Sponsor

PHAC provided input into the study design. PHAC and Mitacs did not have any role in the conduct of the study; collection, management, analysis, and interpretation of the data; preparation, review, or approval of the manuscript; and decision to submit the manuscript for publication.

## Group Authorship

IMPACT Investigators on this project: Natalie Bridger, Janeway Children’s Health C Rehabilitation Centre, St. John’s, NL; Jeannette Comeau, IWK Health Centre, Halifax, NS; Roseline Thibeault, Centre Mere-Enfant de Quebec, CHUL, Quebec City, QC; Dorothy Moore, Jesse Papenburg, The Montreal Children’s Hospital, Montreal, QC; Marc Lebel, Fatima Kakkar, Centre hospitalier universitaire Sainte-Justine, Montreal, QC; Nicole Le Saux, Children’s Hospital of Eastern Ontario, Ottawa, ON; Shaun K. Morris, Kescha Kazmi, The Hospital for Sick Children, Toronto, ON; Rupeena Purewal, Rupesh Chawla, Jim Pattison Children’s Hospital, Saskatoon, SK; Taj Jadavji, Cora Constantinescu, Alberta Children’s Hospital, Calgary, AB; Karina A. Top, Catherine Burton, Stollery Children’s Hospital, Edmonton, AB; Julie A. Bettinger, Laura Sauvé, Manish Sadarangani, BC Children’s Hospital, Vancouver, BC; Jared Bullard, Joanne Embree, Winnipeg Children’s Hospital Health Sciences Center, Winnipeg MB; Jeffrey Pernica, McMaster’s Children Hospital, Hamilton, ON.

## Additional Contributions

We acknowledge all past IMPACT site investigators and RSV working group members involved in IMPACT at the time of data collection. We acknowledge the IMPACT nurse monitors and the staff of the data center.

## Notes

### Author Declarations

Ethics committee/IRB of Alberta Children's Hospital gave ethical approval for this work. Ethics committee/IRB of B.C. Children's Hospital gave ethical approval for this work. Ethics committee/IRB of Le Centre Mere-Enfant de Quebec City gave ethical approval for this work. Ethics committee/IRB of Children's Hospital of Eastern Ontario gave ethical approval for this work. Ethics committee/IRB of CHU-Sainte-Justine Montreal gave ethical approval for this work. Ethics committee/IRB of IWK Health Centre gave ethical approval for this work. Ethics committee/IRB of Eastern Health Janeway Child Health and Rehabilitation Centre gave ethical approval for this work. Ethics committee/IRB of The Montreal Children's Hospital gave ethical approval for this work. Ethics committee/IRB of Jim Pattison Children's Hospital gave ethical approval for this work. Ethics committee/IRB of Stollery Children's Hospital gave ethical approval for this work. Ethics committee/IRB of The Hospital for Sick Children gave ethical approval for this work. Ethics committee/IRB of Children's Hospital, Winnipeg, MB gave ethical approval for this work. Ethics committee/IRB of McMaster's Children Hospital gave ethical approval for this work.

