## Supplemental Material for "Changes in Pediatric RSV Hospitalizations after the COVID-19 Pandemic, 2022-2023, Canada: An Active Surveillance Study"

**Supplemental Table 1.** Counts of Hospitalizations with Respiratory Syncytial Virus (RSV) Detected, Canadian Immunization Monitoring Program, ACTive

**Supplemental Table 2.** Intensive Care Unit Admission and Hospital Length of Stay Longer than 7 days, by Age Group, among Respiratory Syncytial Virus (RSV) Hospitalizations, Canadian Immunization Monitoring Program, ACTive

**Supplemental Table 1.** Counts of Hospitalizations with Respiratory Syncytial Virus (RSV) Detected, Canadian Immunization Monitoring Program, ACTive

| | Number of RSV hospitalizations | |
|---|---|---|
| **Province** | **2017-2018 to 2019-2020** | **2022-2023** |
| **Alberta** | 1480 | 1566 |
| **British Columbia** | 507 | 374 |
| **Manitoba** | 659 | 445 |
| **Newfoundland and Labrador** | 193 | 84 |
| **Nova Scotia** | 452 | 158 |
| **Ontario**[a] | 1429 | 847 |
| **Quebec** | 2449 | 1641 |
| **Saskatchewan** | 380 | 247 |
| **Total** | 7549 | 5362 |

[a] An additional Ontario site participated as of July 1, 2020. Data from this site were excluded from this analysis.

**Supplemental Table 2.** Intensive Care Unit Admission and Hospital Length of Stay Longer than 7 days, by Age Group, among Respiratory Syncytial Virus (RSV) Hospitalizations, Canadian Immunization Monitoring Program, ACTive

| Severity Outcome by age group | (RSV hospitalizations, No. (%) [95%CI])[a] | | | |
|---|---|---|---|---|
| | 2017-2018 to 2019-2020 | 2022-2023 | Difference [95% CI], percentage points[b] | P-value[c] |
| **0-5 months** | | | | |
| **ICU Admission** | 1070 (29) [27.5-30.5] | 782 (35.1) [33.2-37.2] | 6.2 [3.7-8.6] | <0.001 |
| **LOS ≥ 7 days** | 832 (22.5) [21.2-23.9] | 580 (26.1) [24.3-27.9] | 3.5 [1.3-5.8] | 0.024 |
| **6-11 months** | | | | |
| **ICU Admission** | 158 (17) [14.7-19.6] | 108 (16) [13.4-18.9] | -1.1 [-4.7-2.6] | 1 |
| **LOS ≥ 7 days** | 176 (19) [16.6-21.6] | 88 (13) [10.7-15.7] | -6 [-9.5--2.4] | 0.017 |
| **12-23 months** | | | | |
| **ICU Admission** | 230 (17.3) [15.4-19.4] | 133 (14.4) [12.3-16.9] | -2.9 [-5.9-0.2] | 0.845 |
| **LOS ≥ 7 days** | 213 (16) [14.1-18.1] | 101 (11) [9.1-13.1] | -5 [-7.9--2.2] | 0.008 |
| **2-4 years** | | | | |
| **ICU Admission** | 192 (16.4) [14.4-18.6] | 150 (13.2) [11.3-15.2] | -3.3 [-6.1--0.4] | 0.333 |
| **LOS ≥ 7 days** | 253 (21.6) [19.4-24.1] | 172 (15.1) [13.1-17.3] | -6.5 [-9.7--3.4] | <0.001 |
| **5-9 years** | | | | |
| **ICU Admission** | 78 (26.7) [22-32.1] | 61 (20.1) [16-25] | -6.6 [-13.4-0.2] | 0.695 |
| **LOS ≥ 7 days** | 114 (39) [33.6-44.7] | 73 (24.1) [19.6-29.2] | -14.9 [-22.3--7.6] | 0.001 |
| **10-16 years** | | | | |
| **ICU Admission** | 41 (30.1) [23.1-38.3] | 26 (27.1) [19.2-36.7] | -3.1 [-14.8-8.7] | 1 |
| **LOS ≥ 7 days** | 62 (45.6) [37.5-54] | 41 (42.7) [33.3-52.7] | -2.9 [-15.8-10.1] | 1 |

Abbreviations: ICU, intensive care unit; LOS, length of stay; CI, confidence interval

[a] Percentages are calculated among total admissions with RSV for that time period

[b] The percentage point difference was calculated between the 2022-2023 season and the total of the three pre-pandemic seasons, 2017-2018 to 2019-2020. An additional Ontario site participated as of July 1, 2020, but data from this site were excluded from this analysis.

[c] Bonferroni corrected p-value
